# Genomic epidemiology of the rotavirus G2P[4] strains in coastal Kenya pre- and post-rotavirus vaccine introduction, 2012 – 2018

**DOI:** 10.1101/2022.10.21.22281210

**Authors:** Timothy O. Makori, Joel L. Bargul, Arnold W. Lambisia, Mike J. Mwanga, Nickson Murunga, Zaydah R. de Laurent, Clement S. Lewa, Martin Mutunga, Paul Kellam, Matthew Cotten, D. James Nokes, My Phan, Charles N. Agoti

## Abstract

The introduction of rotavirus vaccines into the national immunization programme in many countries has led to a decline of childhood diarrhoea disease burden. Coincidentally, the incidence of some rotavirus group A (RVA) genotypes, has increased, which may result from non-vaccine-type replacement. Here we investigate the evolutionary genomics of rotavirus G2P[4] which has shown an increase in countries that introduced the monovalent Rotarix® vaccine. We examined the 63 RVA G2P[4] strains sampled from children (aged below 13 years) admitted to Kilifi County Hospital, Coastal Kenya, pre- (2012 to June 2014) and post- (July 2014-2018) rotavirus vaccine introduction. All the 63 genome sequences showed a typical DS-1 like genome constellation G2-P[4]-I2-R2-C2-M2-A2-N2-T2-E2-H2. G2 sub-lineage IVa-3 strains predominated in the pre-vaccine era co-circulating with low numbers of G2 sub-lineage IVa-1 strains, whereas sub-lineage IVa-3 strains dominated the post-vaccine period. In addition, in the pre-vaccine period, P[4] sub-lineage IVa strains co-circulated with low numbers of P[4] lineage II strains, but P[4] sub-lineage IVa strains predominated in the post-vaccine period. On the global phylogeny, the Kenyan pre- and post-vaccine G2P[4] strains clustered separately, suggesting that different virus populations circulated in the two periods. However, the strains from both periods exhibited conserved amino acid changes in the known antigenic epitopes, suggesting that replacement of the predominant G2P[4] cluster was unlikely a result of immune escape. Our findings demonstrate that the pre- and post-vaccine G2P[4] strains circulating in Kilifi, coastal Kenya, differed genetically, but likely were antigenically similar. This information informs the discussion on the consequences of rotavirus vaccination on rotavirus diversity.

## INTRODUCTION

In 2009, the World Health Organization (WHO) recommended inclusion of rotavirus vaccines into the national immunization programmes (NIPs) globally [1]. Kenya introduced the WHO pre-qualified Rotarix® RVA (rotavirus group A) vaccine into its NIP in July 2014 [2]. With the increasing uptake of rotavirus vaccines globally, there has been a significant reduction in RVA-associated disease burden, but this virus still caused about 128,500 deaths in 2016 alone [3,4], with the majority of cases occurring in low-income countries [5]. In Kenya, post-vaccine introduction impact studies have reported significant reduction of rotavirus-associated diarrhoea hospitalization in children under five years, a vaccine effectiveness of ∼64% [6], and a significant increase in the positivity rate of Rotarix® heterotypic genotypes such as G2P[4] and G3P[8] [7,8]. Similar findings were observed in other countries that introduced the Rotarix® vaccine in their NIPs [9–18]

Whole genome analysis of RVA can reveal the transmission and evolutionary history of circulating strains, including emerging mutations, and the origins and genetic diversity of the strains circulating in a particular region [19]. The rotavirus G2P[4] genotype is known to possess a DS-1 like genomic constellation (G2-P[4]-I2-R2-C2-M2-A2-N2-T2-E2-H2) [20]. G2P[4] strains are believed to have undergone genetic evolution in a stepwise pattern [21]. This is from lineage I to IVa in the NSP5, NSP1, VP2, VP4, and VP7 genome segments, and from lineage I to V in the VP1, VP3, VP6, NSP2, NSP3, and NSP4 genome segments, with some of the strains undergoing intragenotype reassortments in the VP7, VP3, and NSP4 genes after 2004 giving rise to emergent lineages of V in the VP7 segment, lineages VI and VII in the VP3 gene, and VI, VII, VIII, IX and X lineages in the NSP4 gene [21–23]. The G2P[4] lineages circulating in Kenya are unknown.

Characterization of South African G2P[4] strains, through comparison of strains occurring during pre-and post-introduction of Rotarix® vaccine, revealed sub-lineage shifts from G2 sub-lineage IVa-1 to G2 IVa-3, and P[4] sub-lineage IVa to P[4] IVb, and these shifts in genetic evolution were attributed to arise due to natural fluctuations and not as a result of vaccine pressure [24]. Similarly, G2P[4] whole genome analysis in Ghana [22], Australia [25,26], South Korea [11], Bangladesh [27], and Brazil [28] showed that implementation of Rotarix® vaccination does not influence genetic diversity of the circulating G2P[4] strains and common amino acid replacements in the VP7 antigenic epitopes including A87T, D96N, S213D were reported, irrespective of the vaccination period.

The main goal of this study was to conduct whole genome analysis of the G2P[4] genotypes circulating in Kilifi County, Kenya, to determine whether introduction of Rotarix® vaccine in Kenya’s NIP impacts on the genetic diversity of the circulating G2P[4] viral populations. Data from this study were compared with contemporaneous global RVA strains to establish the phylogenetic context and potential origin of the Kenya’s pre- and post-vaccination rotavirus G2P[4] strains.

## MATERIALS AND METHODS

### Ethical approval

We collected samples from children to screen for and determine the genetic diversity of rotavirus group A (RVA) viral populations. The study was conducted with strict adherence to the study protocols approved by the Scientific and Ethics Review Unit (REF: #3049 and #2861) at the Kenya Medical Research Institute, KEMRI, Nairobi, Kenya. Parents and guardians of the eligible children were provided with sufficient information about the research study to allow each individual to make informed and independent decisions for their children to be enrolled in the study. Sample collection proceeded after obtaining written informed consent from the parents and guardians.

### Study participants

The study was based at Kilifi County Hospital (KCH), a referral health facility that mainly serves the people of Kilifi County located in the North Coast of Kenya [29]. Stool samples were obtained from children, aged below 13 years, who presented with diarrhoea as one of the illness symptoms and were admitted at KCH [6,30]. Diarrhoea was defined as passing of watery stool at least three times in the last 24 hours.

The stool samples were screened to detect RVA by using ELISA kit (ProSPect™; Oxoid, Basingstoke UK). All the RVA positive samples were initially genotyped by partial segment sequencing approach [7]. VP7 and VP4 genes were sequenced, and G and P genotypes inferred using the Virus Pathogen Resource (VIPR) tool for RVA [31]. Those classified as G2P[4] based on the outer capsid proteins were selected for this study (Fig. 1) [7].

**Fig. 1:**
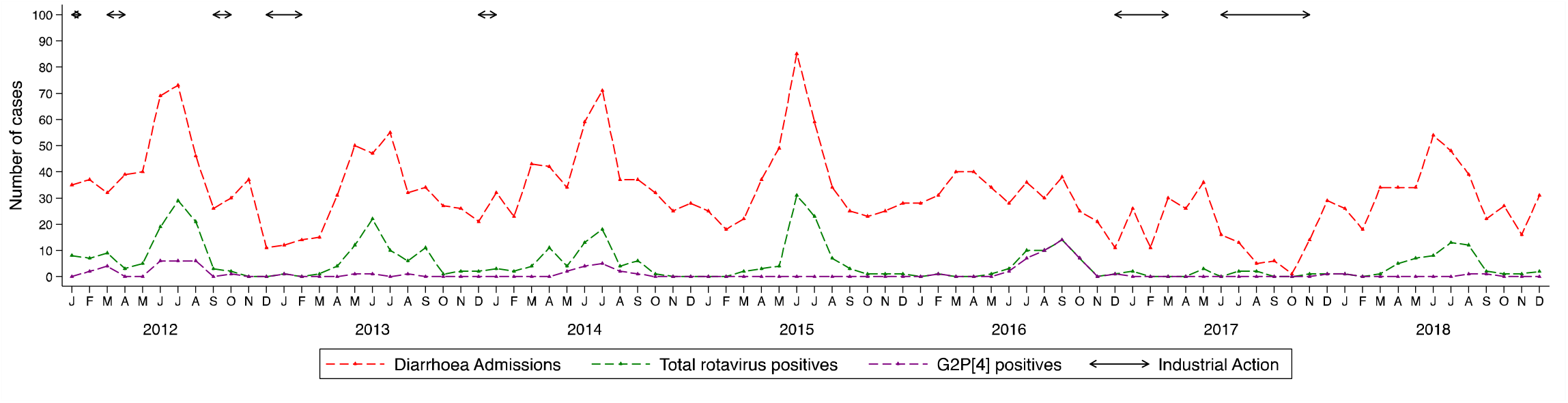
Temporal pattern of cases of diarrhoea, rotavirus A and G2P[4] genotype in Kilifi County Hospital between 2012 and 2018. The red line shows monthly diarrhoea admissions, the green line shows the monthly RVA positives, the purple line shows the monthly G2P[4] detected during the study period, while the topmost solid lines show periods in which there were healthcare industrial actions at KCH. Determination of the G and P genotypes was by partial segment sequencing using the Sanger approach [7].

### RNA extraction and processing

The samples collected in the pre-vaccine period (January 2012 – June 2014) were sequenced using agnostic whole genome sequencing method [32], while the post-vaccine samples (July 2014 – December 2018) were sequenced using an amplicon-based whole genome sequencing approach [33,34].

Processing of pre-vaccine samples was done by centrifugation of 110 μL of stool suspension in PBS (phosphate buffered saline) for 10 minutes at 10,000 × g. Next, 2 U/μl of TURBO DNase (#AM2238; Life Technologies, Carlsbad, USA) were added to degrade non-encapsulated DNA. Nucleic acid extraction was performed according to the Boom method [35]. First strand cDNA was synthesized using the SuperScript III Reverse Transcriptase Kit (#18064014; Life Technologies, Carlsbad, USA) with non-ribosomal random hexamer primers [36]. Second strand cDNA was synthesized using 5U of Klenow fragment 3’ – 5’ exo-(#M0212S; New England Biolabs, Ipswich, USA).

Processing of post-vaccine introduction samples was done by subjecting 200 mg of stool specimens to bead beating [37], followed by nucleic acid extraction using the QIAamp Fast DNA Stool Mini kit (#51604; Qiagen, Manchester, UK) following the manufacturer’s instructions. Reverse Transcription-PCR (RT-PCR) was performed using the SuperScript IV One-Step RT-PCR System (#2594025, Thermo Fisher Scientific, Waltham, USA) following the manufacturer’s instructions. The published primers used in our study to conduct RT-PCR assays were adopted from earlier studies (Table S1) [33,34]. The PCR conditions for the non-structural gene segments (NSP1, NSP2, NSP3, NSP4, and NSP5) consisted of 40 cycles of thermocycling (30 seconds at 90°C, one minute at 55°C, and four minutes at 68°C), whereas amplification of structural gene segments (VP1, VP2, VP3, VP4, VP6, and VP7) included 40 cycles of thermocycling (90°C for 30 seconds, 61°C for one minute and 68°C for six minutes), and included a final extension at 72°C for four minutes. PCR amplicons were resolved under a 2% agarose gel stained with RedSafe (iNtRON Biotechnology, Inc) for visualization of DNA bands. PCR products were purified using Exonuclease I (#EN0581; Thermo Fisher Scientific, Waltham, USA) as described by the manufacturer and pooled for each sample.

### Next generation sequencing

Preparation of standard Illumina libraries for pre-vaccine samples was performed according to the published protocol [32]. Briefly, the double-stranded cDNA for each sample was sheared to obtain 400 – 500 nucleotide fragments. Each sample was then indexed separately to unique adapters and multiplexed at 95 samples and then sequenced on a HiSeq platform to generate about 1.5 million 250bp paired end reads per sample.

For the post-vaccine samples was done by purifying the pooled amplicons for each sample using the Agencourt AMPure XP Kit (#A63881; Beckman Coulter, USA) as described by the manufacturer. Library preparation was performed using the Illumina DNA flex (#20025519, Illumina, San Diego, USA) as per the manufacturer’s specifications. Briefly, bead-linked transposomes were used to tagment the DNA, followed by addition of adapters to the DNA fragments using a limited PCR program. The adapter-linked DNA was cleaned using the tagment wash buffer. After that, the purified tagmented DNA was amplified via a limited-cycle PCR program that adds the i7, i5 adapters and sequences required for cluster generation during sequencing. Next, the amplified libraries were purified using a double-sided bead purification method. Subsequently, each DNA library was quantitated, and correct insert sizes confirmed on an Agilent 2100 Bioanalyzer using the Agilent high sensitivity DNA kit (#5067; Agilent, Santa Clara, USA). The DNA libraries were quantified on the Qubit fluorimeter 2.0 using the Qubit dsDNA HS Assay kit (#Q32851, Life Technologies, Thermo Fisher Scientific, Waltham, USA), normalized, and pooled at equimolar concentrations. Pooled DNA libraries were denatured and sequenced on the Illumina MiSeq platform (Illumina, San Diego, USA) to generate 150 paired end reads.

### Genome assembly

Quality trimming of Illumina FASTQ reads was done using Trimmomatic (Phred score >30) with the following flags “ILLUMINACLIP: adapters_file: 2:30:10 LEADING:3 TRAILING:3 SLIDINGWINDOW: 4:15 MINLEN:36” to remove adapters and low-quality bases [38]. De-Novo assembly of the quality trimmed reads was done using Spades with the following flags “-k 99,127 --careful” [39]. For the pre-vaccine sequences, RVA-specific contigs were identified using USEARCH [40] and a SLIM algorithm [41]. Partial and overlapping contigs were joined using Sequencher [42] to obtain full-length sequences. For the post-vaccine sequences, Quast was used to check the quality of the contigs [43]. Next, artemis was employed to determine the ORFs of each RVA segment [44]. Then, genotyping of the assembled pre- and post-vaccine sequences was done using the Virus Pathogen Resource tool for RVA [31]. The nucleotide sequences generated in this study have been deposited into GenBank under accession numbers MZ093788 to MZ097268 and OP677569 to OP677754 (Table S2).

### Global sequences collection and processing

All available G2P[4] sequences irrespective of the sequence length and their corresponding metadata, including year of collection and location were downloaded from the Virus Pathogen Resource of RVA [31]. Records missing metadata were manually searched and any information, including location and collection year, that could be found in the primary publications was included in the respective sequence data. The sequences were subset to obtain datasets of each genome segment. The datasets of all the genome segments were filtered to only include samples with all the 11 segments. For all the 11 segments, more that 80 % of the coding sequence (CDS) region was considered for analysis. Overall, 350 global sequences for each segment met the inclusion criteria for phylogenetic analyses (Table S2).

### Phylogenetic analysis

The global dataset was combined with the sequences of this study for each genome segment and aligned using MAFFT (v7.487) with the command “mafft --auto --reorder --preservecase input_file.fasta > output_file.fasta” [45]. Maximum likelihood phylogenetic trees were reconstructed using IQTREE2 (v2.1.3) [46] using the best model selection [47] and 1000 bootstrap replication settings [48]. The ML trees were linked to the respective metadata in R v4.1.0 and the “ggTree” R package used to plot and visualize the trees [45]. For lineage designation, sequences of previously described lineages for each segment [21–23] were utilized as the references (Table S2).

### Statistical analyses

All statistical analyses were carried out using Stata v13.1 [49]. Chi-square test was used to compare among groups, with P< 0.05 indicating statistical significance.

## RESULTS

### Baseline characteristics of the study participants

Peaks of rotavirus A (RVA) infections coincided with cases of diarrhoea during the study period between May and September (Fig. 1). However, sample collection was majorly disrupted in 2016 and 2017 by strikes of health care providers (Fig. 1) [6]. One huge peak of G2P[4] infections was observed between June and November 2016 (post-vaccine) and a small one documented between May and September 2012 (pre-vaccine) (Fig. 1).

No significant difference was reported in age, age groups, gender, and vaccination status between the cases infected with G2P[4] genotypes and the non-G2P[4] genotypes (P>0.05) (Table 1). G2P[4] genotypes were detected in 87 samples (20.3%), of which 63 (14.7%) were successfully full-genome sequenced (Table 1). Of the sequenced samples, 13 (20.6%) were from children who received two doses of the Rotarix® vaccine (Table 1).

**Table 1:** Baseline demographic characteristics of the children that were rotavirus positive and those found to be G2P[4] infected during the study period (2012 to 2018)

| Characteristics | RVA<br>Positives<br>(%) | Successfully<br>Sequenced<br>G2P[4]<br>(%) | All<br>G2P[4]<br>(%) | non-G2P [4]<br>(%) | All G2P[4] vs<br>non-G2P[4] P-value |
| --- | --- | --- | --- | --- | --- |
| Total cases | 429 | 63 (14.7) | 87 (20.3) | 342 (79.7) |  |
| Vaccine period |  |  |  |  | 0.039 |
| Pre-vaccine | 215 (49.9) | 33 (52.4) | 35 (40.2) | 180 (52.6) |  |
| Post-vaccine | 214 (50.1) | 30 (47.6) | 52 (59.8) | 162 (47.4) |  |
| Age (in months) |  |  |  |  | 0.944 |
| Mean (SD) | 14.9 (13.1) | 17.6 (16.4) | 16.9 (15.4) | 14.4 (12.4) |  |
| Median (IQR) | 11.7 (8.3-17.9) | 11.2 (8.6-20.7) | 10.6 (8.4-19.7) | 12.2 (8.3-16.8) |  |
| Age group (in<br>months) |  |  |  |  | 0.092 |
| 0-11m | 216 (50.4) | 33 (52.4) | 47 (54.0) | 216 (50.4) |  |
| 12– 23m | 169 (39.4) | 20 (31.2) | 26 (29.9) | 143 (41.8) |  |
| 24– 59m | 35 (8.2) | 7 (11.1) | 11 (12.6) | 24 (7.0) |  |
| >=60 m | 9 (2.1) | 3 (4.8) | 3 (3.5) | 6 (1.7) |  |
| Gender |  |  |  |  | 0.789 |
| Male | 251 (51.5) | 37 (58.7) | 52 (59.7) | 199 (58.2) |  |
| Vaccination<br>status |  |  |  |  | 0.129 |
| Vaccinated | 88 (20.6) | 14 (22.2) | 22 (25.6) | 66 (19.4) |  |
| Not vaccinated | 215 (50.4) | 33 (52.4) | 35 (40.7) | 180 (52.8) |  |
| Unknown | 124 (29.0) | 16 (25.4) | 29 (33.7) | 95 (27.9) |  |
| <sup>1</sup>Full vaccination | 79 (18.4) | 13 (20.6) | 21 (24.1) | 58 (17.0) | 0.256 |

<sup>1</sup> Full vaccination implies that the participants received two doses of the Rotarix vaccine as recommended by WHO

### Genome constellations

To determine the genetic diversity in the Kilifi G2P[4] strains and their genetic relatedness with global strains, near full-genome sequences (>80% genome coverage) of 63 Kilifi samples were sequenced from the pre- (n=33) and the post- (n=30) vaccine periods (Table S2). Using the Virus Pathogen Resource (VIPR) for RVA genotype determination [31], all the 63 sequences were classified as G2-P[4]-I2-R2-C2-M1-A2-N2-T2-E2-H1 genotype (DS-1-like typical genome constellation) as shown in Table S3.

### Phylogenetic and sequence analysis

To gain insights into the genetic diversity of the study G2P[4] strains in the global context, genetic distance-resolved phylogenetic trees were constructed for all the 11 gene segments (Fig. 2 & Fig. S1). Sequence identity matrices of the study G2P[4] strains exhibited high nucleotide sequence similarities (93 – 100%) in the NSP1, NSP2, NSP3, NSP5, VP1, VP2, VP4, VP6, and VP7 genome segments, and low to high nucleotide sequence similarity (85 – 100%) in the VP3 and NSP4 genes (Table 2).

**Table 2:** Percentage nucleotide and amino acid identity for the Kilifi G2P[4] strains

| Genome<br>segment | Percentage<br>nucleotide<br>similarity | Percentage<br>amino<br>acid<br>similarity |
| --- | --- | --- |
| VP4 | 94.3-100 | 96.8-100 |
| VP7 | 94.3-100 | 96.3-100 |
| VP6 | 94.0-100 | 96.0-100 |
| VP1 | 93.9-100 | 98.0-100 |
| VP2 | 97.2-100 | 99.2-100 |
| VP3 | 87.1-100 | 92.1-100 |
| NSP1 | 95.7-100 | 96.0-100 |
| NSP2 | 97.3-100 | 97.5-100 |
| NSP3 | 97.1-100 | 98.4-100 |
| NSP4 | 85.0-100 | 91.5-100 |
| NSP5 | 94.5-100 | 94.5-100 |

**Fig. 2:**
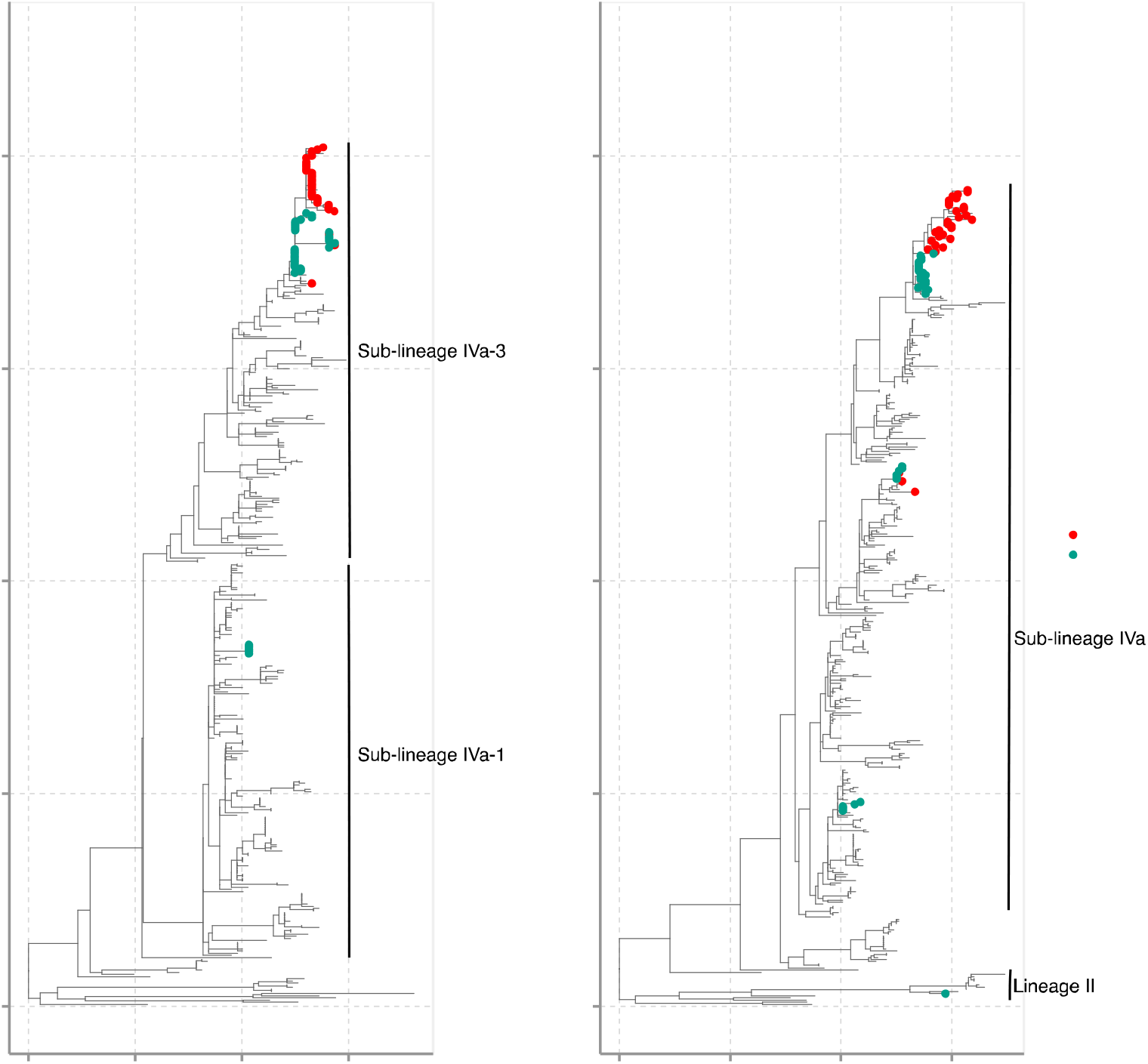
Phylogenetic reconstruction of the 63 Kilifi G2P[4] sequences against a backdrop of 350 global sequences for the VP7 and VP4 RVA genome segments using maximum likelihood (ML) methods. The Kilifi sequences are colored by the period of sample collection (either before or after vaccine introduction in Kenya). For all the global sequences, more than 80% of the coding sequence (CDS) region was considered for analysis. For the Kilifi sequences, >80% CDS were used for the VP7 and 68% for the VP4 segment. The study sequences classified to the lineages indicated in each phylogenetic tree.

### Analysis of the VP7 gene

The VP7 gene is highly variable and encodes the humoral immune response glycoprotein [50]. The VP7 genetic distance-resolved phylogenetic tree showed that the Kilifi sequences formed three clusters: a monophyletic cluster, a minor monophyletic cluster, and a singleton (Fig. 2). Within the major cluster, the Kilifi strains separated by vaccination period, with one subcluster consisting of strains circulating two years after Rotarix® vaccine introduction and were interspersed with three strains isolated from children admitted to Kenyatta National Hospital (KNH), Kenya in 2017 (Fig. 2). These sequences shared two non-synonymous amino acid substitutions (S72G, S75L) with respect to the pre-vaccine strains (Table S4). The second subcluster mainly consisted of strains circulating in the pre-vaccine period and two strains that circulated in July 2014, i.e., early post-vaccine period (Fig. 2). The sequences in the minor monophyletic cluster consisted of five Kilifi strains collected in 2012, while the singleton Kilifi strain (KLF1033/2018) clustered with three strains detected in Mozambique in 2013 (Fig. 2).

With regards to the VP7 lineages, the Kilifi G2 strains were classified into lineage IV and further classified into sub-lineage IVa-1 and IVa-3 (Fig. 2). In 2012, sub-lineages IVa-1 and IVa-3 sequences co-circulated in Kilifi, while in the global context sub-lineages IVa-1, IVa-3 and IV non-a co-circulated (Fig. 3A). However, IVa-1 strains in Kilifi were replaced with sub-lineage IVa-3 strains in 2013 that dominated until 2018 (Fig. 3A), unlike in the global context where sub-lineages IVa-1, IVa-3 and V co-circulated in 2013, sub-lineage IVa-1 predominantly circulated in 2014, IVa-1 and V co-circulated in 2015, and IVa-3 re-emerged in 2016 replacing lineage V and co-circulated with sub-lineage IVa-1 until 2018 (Fig. 3A). No lineage shift was observed pre- and post-vaccine introduction.

**Fig. 3:**
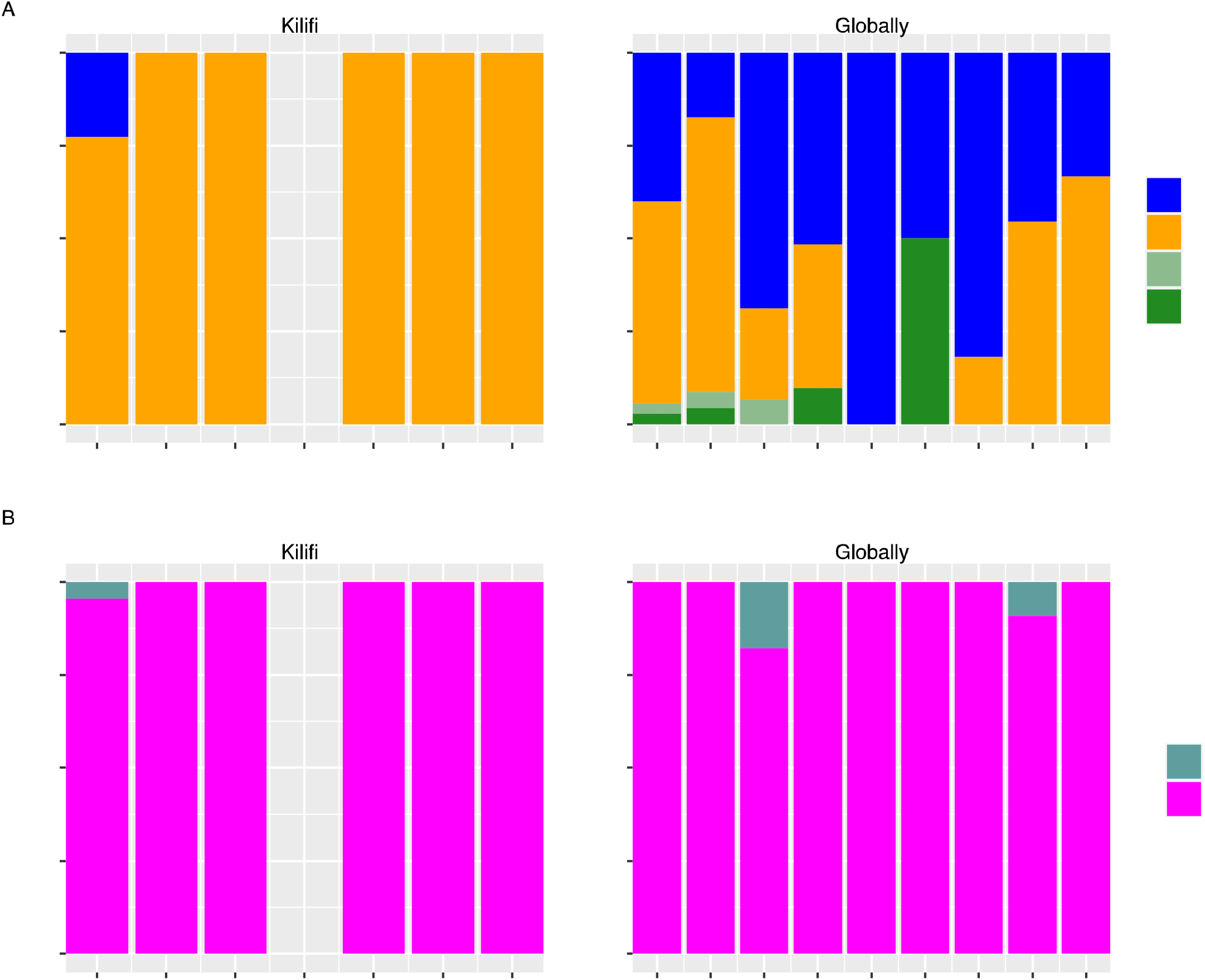
Temporal pattern of the G2 and P[4] lineages observed in Kilifi and globally. (A) Temporal pattern of the Kilifi G2 lineages from 2012 to 2018 and temporal pattern of the global G2 lineages from 2010 to 2018. (C) Temporal pattern of the Kilifi P[4] lineages from 2012 to 2018 and temporal pattern of the global P[4] lineages from 2010 to 2018.

### Analysis of the VP4 gene

The VP4 gene is highly variable and encodes a highly immunogenic protease sensitive protein involved in receptor binding and cell penetration [50]. In the VP4 phylogenetic tree, the P[4] Kilifi sequences formed clusters (n>2) mainly based on the vaccination period separated from global sequences (Fig. 2). However, two Kilifi sequences formed singletons, with the KLF1033/2018 strain clustering with a sequence isolated from a child admitted to KNH, while the KLF0616/2012 strain was interspersed with sequences from Mozambique (Fig. 2). A major cluster of Kilifi sequences further sub-clustered based on the vaccination period, with the post-vaccine sequences interspersing with Kenyan sequences isolated from children admitted to KNH (Fig. 2). In addition, Kilifi strains collected in 2014 and some 2012 strains formed two distinct clades, clustering separately from global sequences (Fig. 2).

During the study period, G2 lineages II and IVa strains circulated in Kilifi, with lineages II and IV co-circulating in 2012, consistent with the global context (Fig. 3B). From 2013 to 2018, sub-lineage IVa predominantly circulated in Kilifi similar the global context (Fig. 3B). However, in the global context, few lineage II strains co-circulated with IVa strains in 2017 (Fig. 3B). No lineage shift was observed pre- and post-vaccine introduction (Fig 2).

### Analysis of the backbone genome segments

The backbone genome segments of the Kilifi G2P[4] strains (VP6, VP1-VP3, and NSP1-NSP5) formed up to four clusters on the global phylogenetic trees (Fig.S1). In the VP6, VP1, VP2, VP3, NSP1, and NSP2 genes, majority of the Kilifi sequences formed one major cluster which further separated into two sub-clusters of only pre- and post-vaccine sequences (Fig. S1). The post-vaccine strains in these genes clustered closely with 2017 sequences from KNH, Kenya (Fig.S1). In addition, the Kilifi 2014 sequences in the VP6, VP3, NSP1, and NSP2 segments exhibited a different clustering pattern of a further minor sub-cluster irrespective of the vaccination period, consistent with the VP4 gene (Fig. S1). Four post-vaccine sequences were interspersed with the pre-vaccine sequences in the NSP4 gene and a singleton of post-vaccine sequence clustered with pre-vaccine sequences in the NSP3 gene (Fig. S1). The NSP5 post-vaccine sequences formed one cluster, while the pre-vaccine sequences exhibited a different clustering pattern consisting of three distinct clusters (n≥2) and three singletons (KLF0601/2012 was interspersed with a sequence from KNH, KLF1066/2014 and KLF0722/2014) (Fig. S1). A minor cluster consisting of five Kilifi pre-vaccine strains (KLF0550/2012, KLF0551/2012, KLF0553/2012, KLF0558/2012, and KLF1064/2012) separate from global sequences was observed in the VP1, VP2, VP3, NSP2, NSP4, and NSP5 genes; however, interspersed with sequences that circulated between 2012 and 2017 in Japan, Hungary, Australia, and Belgium in the NSP3 gene and further formed two sub-clades in the VP6 gene (Fig. S1). In addition, a minor cluster of Kilifi pre-vaccine sequences (KLF0640/2013, KLF0673/2013, and KLF0657/2013) interspersed with sequences from Malawi was observed in the VP3 gene, indicating possible importation of these strains (Fig. S1). Besides, a singleton of a post-vaccine sequence (KLF1033/2018) was interspersed with sequences from Mozambique that circulated in 2013 across all the backbone genes (Fig. S1). For the NSP5 gene, three other singletons were observed: KLF0601/2012 interspersed with a sequence from KNH, KLF1066/2014 and KLF0722/2014 each separate from global sequences (Fig. S1). No lineage shifts were reported in the backbone genome segments (Fig. S1).

### Amino acid changes in the VP7 glycoprotein (G) and VP4 Protease sensitive (P) proteins

The pattern of amino acid (aa) substitutions in the G and P proteins was analysed in relation to the ancestral DS-1 sequence. The VP7 gene contains 7-1 (7-1a and 7-1b) and 7-2 antigenic epitopes, which affect the ability of antibodies to neutralize virus infectivity and reduce vaccine effectiveness [51]. The Kilifi strains exhibited 17 aa changes relative to the DS-1 VP7 ancestral sequence (Table S4). All the Kilifi strains had three aa mutations (D96N, N125T, and V129M) in the 7-1a antigenic epitope with respect to the DS-1 ancestral strain (Fig. 4 & Table S4). Furthermore, except for the 2014 sequences, the Kilifi sequences exhibited an A87T aa mutation in the 7-1a epitope (Table S4). Compared to the DS-1 sequence, the Kilifi sub-lineage IVa-1 strains exhibited the N242 aa mutation, whereas sub-lineage IVa-3 strains harboured the N213D aa mutation; both in the 71-b epitope. (Table S4). Besides, the I44M aa change was observed in the T lymphocyte epitope (40 – 52) of all the Kilifi strains (Table S4).

**Fig. 4:**
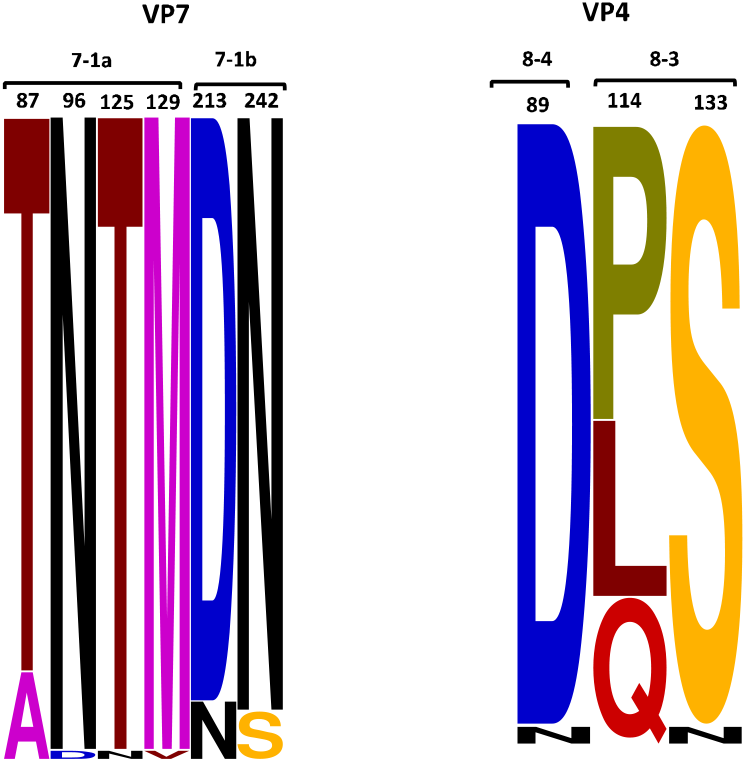
A sequence logo showing the amino changes observed in known antigenic epitopes of G2 and P[4] proteins in the Kilifi G2P[4] relative to the ancestral DS-1 strain. The changes in amino acids are shown in different color schemes.

The VP4 surface protein is cleaved into the VP8* and the VP5* domains containing the 8-1 to 8-4 and 5-1 to 5-5 antigenic epitopes [52]. Analysis of the VP4 aa mutations revealed that the Kilifi strains differed only at three positions; Q114P or L114P and N133S in the 8-3 epitope and N89D in the 8-4 epitope, relative to the DS-1 prototype sequence (Fig 4 & Table S5).

## DISCUSSION

We investigated the evolutionary dynamics of G2P[4] strains sampled from children admitted to Kilifi County Hospital, Coastal Kenya between 2012 and 2018. All the recovered genomes showed a typical DS-1 like constellation consistent with findings from several countries using the Rotarix® vaccine in their NIPs [21,22,24,53].

Separate clusters of pre- and post-vaccine sequences were observed in the VP1-VP4, VP6, VP7, NSP1, NSP2, and NSP5 segments. However, the strains sampled in July 2014 (early post-vaccine period) clustered with pre-vaccine strains across all the 11 gene segments in our study possibly because the vaccine coverage was low and thus had no impact yet on the circulating genotypes and lineages. Unique clusters of either G2P[4] or G1[P8] strains separated by RVA vaccine period have been reported in South Africa [24], Rwanda [54], Australia and Belgium [55], which were interpreting as reflecting natural genetic fluctuations rather than vaccine induced evolution. However, the study NSP4 and NSP3 genes exhibited some clusters of mixed pre- and post-vaccine sequences, indicating that in some strains the pre-vaccine may have reassorted with post-vaccine introduction strains as similarly suspected in Mozambique, albeit in G1P[8] [56].

Phylogenetic analyses indicated that the diversity of the Kilifi G2P[4] strains may have been locally restricted both in the pre- and post-vaccine periods, since Kilifi sequences clustered away from global sequences in all the 11 genome segments. In addition, limited sequence data from Kenya and East Africa may have contributed to this uncertainty about the regional context of Kilifi diversity. The Kilifi post-vaccine strains only clustered with Kenyan sequences sampled from children admitted to Kenyatta National Hospital in 2017, further suggesting locally restricted genetic evolution. However, one sample (KLF1033/2018) consistently clustered with sequences from Mozambique and some pre-vaccine strains were interspersed with global strains for the NSP3 segment, suggestive of limited introduction from other countries. No lineage shift was observed pre- and post-periods in Kilifi inconsistent with findings in South Africa [24] where RVA vaccine introduction resulted in lineage shift. Furthermore, few G2 and P[4] lineages were in circulation within Kilifi compared with the combined global data during the study period. This supported our hypothesis that local drivers were responsible for the diversity within the Kilifi setting.

The Kilifi strains harbored six conserved amino acid (aa) substitutions in the VP7 antigenic epitopes; 7-1a and 7-1b with respect to the ancestral DS-1 G2P[4] strain. Three of these positions (A87T, D96N, and N213D) are critical for antibody binding and sequence changes here may lead to escape from host neutralizing antibodies [57]. The I44M aa change may affect cellular immunity as this region harbors a known T lymphocyte epitope (40-52) of the VP7 genes. All Kilifi strains had this change that potentially result in loss of recognition by T cells leading to escape from host immune responses [58,59]. Three aa acid changes were observed in VP4 antigenic epitopes in 8-4 (N89D) and 8-3 (Q114P or L114P and N133S) in the Kilifi strains. These have been associated with escape of attachment of the virus to host neutralizing monoclonal antibodies [60]. These aa substitutions were present in both pre- and post-vaccine Kilifi strains, suggesting they were not brought about by vaccine use.

This study had limitations. First, only sequences sampled from hospitalized children were analysed, thus may not conclusively reflect diversity that was in circulation in the entire coastal Kenya population. We only analysed a few genomes across the years. Second, we only recovered near complete genomes. Only 68% coverage was recovered in the VP4 segment.

In conclusion, our study reinforces the significance of genomic sequencing in monitoring the effect of vaccine pressure on circulating RVA strains in Kenya. The Kilifi strains to a large extent clustered based on the vaccination period and were separate from the global strains. Furthermore, conserved amino acid mutations were observed in the VP7 and VP4 antigenic epitopes of the pre- and post-vaccine strains, suggesting that the Rotarix® vaccine did not have a direct impact on the evolution of the circulating strains.

## Data Availability

All the sequence data produced in the present work are available online under accessions under MZ093788 to MZ097268 and OP677569 to OP677754.
All the epidemiological produced in the present work are available online at Virus Epidemiology and Control Dataverse

https://doi.org/10.7910/DVN/P4MRVF

## Data availability

The epidemiological data is available on the VEC dataverse (https://doi.org/10.7910/DVN/P4MRVF).

## Acknowledgement

We thank all the study participants for their contribution of study samples, their parents/guardians, members of the viral epidemiology and control research group (http://virec-group.org/) and colleagues at the KEMRI Wellcome Trust Research Programme for their useful discussions during the preparation of the manuscript. This paper is published with the permission of the Director of KEMRI.

## Supplementary Figure Legend

Fig. S1: Phylogenetic reconstruction of the 63 Kilifi G2P[4] sequences against a backdrop of 350 global sequences for the VP6, VP1, VP2, VP3, NSP1, NSP2, NSP3, NSP4, and NSP5 RVA genome segments using maximum likelihood (ML) methods. The Kilifi sequences are colored by the period of sample collection (either before or after vaccine introduction in Kenya). For the Kilifi and all the global sequences, more than 80% of the coding sequence (CDS) region was considered for analysis. The study sequences classified to the lineages indicated in each phylogenetic tree.

