## Supplementary material for "Genomic epidemiology of the rotavirus G2P[4] strains in coastal Kenya pre- and post-rotavirus vaccine introduction, 2012 – 2018": Table S1

| Segment | Primer name | 5’ sequence 3’ | Product Size | Full segment size | Source publication |
| --- | --- | --- | --- | --- | --- |
| VP1 | VP1-1F | TGTAAAACGACGGCCAGTGGCTATTAAAGCTGTAC | 3269 | 3302 | Magagula et al. 2015 |
|  | VP1- 3302-R | CAGGAAACAGCTATGACCGGTCACATCTAAGCAC |  |  |  |
| VP2 | VP2-1F | TGTAAAACGACGGCCAGTGGCTATTAAAGGCTCAATG | 2695 | 2717 | Magagula et al. 2015 |
|  | VP2-2723R | CAGGAAACAGCTATGACCGGTCATATCTCCACAGTG |  |  |  |
| VP3 | VP3-1F | TGTAAAACGACGGCCAGTGTTTTACCTCTGATGGTG | 2530 | 2591 | Magagula et al. 2015 |
|  | VP3-2591R | CAGGAAACAGCTATGACCGGTCACATCATGACTAG |  |  |  |
| VP4 | VP4-1F | TGTAAAACGACGGCCAGTGGCTATAAAATGGCTTC | 2333 | 2359 | Magagula et al. 2015 |
|  | VP4-2359R | CAGGAAACAGCTATGACCGGTCACATCCTCAATAG |  |  |  |
| VP6 | VP6-1F | TGTAAAACGACGGCCAGTGGCTTTWAAACGAAGTCTTC | 1320 | 1356 | Magagula et al. 2015 |
|  | VP6-1364R | CAGGAAACAGCTATGACCGGTCACATCCTCTCAC |  |  |  |
| VP7 | VP7-1F | TGTAAAACGACGGCCAGTGGCTTTAAAAGAGAGAATTTC | 1023 | 1059 | Magagula et al. 2015 |
|  | VP7-1063R | CAGGAAACAGCTATGACCGGTCACATCRWACAATTC |  |  |  |
| NSP1 | NSP1 F | GGCTTTTTTTATGAAAAGTCTTGTG | 1547 | 1564 | Fujii et al. 2012 |
|  | NSP1 R | CTAGGCGCTACTCTAGT |  |  |  |
| NSP2 | NSP2 F | GGCTTTTAAAGCGTCTCAGTC | 1058 | 1058 | Fujii et al. 2012 |
|  | NSP2 R | GGTCACATAAGCGCTTTCTATTC |  |  |  |
| NSP3 | NSP3 F | GGCTTTTAATGCTTTTCAGTGGTTG | 1050 | 1050 | Fujii et al. 2012 |
|  | NSP3 R | GGTCACATAACGCCCCTATAG |  |  |  |
| NSP4 | NSP4 F | CTTTTAAAAGTTCTGTTCCGAGAG | 739 | 750 | Fujii et al. 2012 |
|  | NSP4 R | AAGACCATTCCTTCCATTAAC |  |  |  |
| NSP5 | NSP5 F | GGCTTTTAAAGCGCTACAGT | 663 | 663 | Fujii et al. 2012 |
|  | NSP5 R | GGTCACAAAACGGGAGTGGGGA |  |  |  |
| Some degenerate base symbols have been used (R = A or G, W = A or T).  The product size and genome size are based on the Wa strain. | | | | | |

Table S1: RVA segment specific primers used in the one-step RT-PCR
